# Ejaculatory Function and Clinical Outcomes Following Robotic Aquablation for Prostatic Bladder Outflow Obstruction: A Retrospective Real-World Cohort Study Protocol

**DOI:** 10.64898/2026.05.28.26354125

**Authors:** D Shroff, T Newman, S Malde, C Martyn-Hemphill

**Affiliations:** Guy’s Hospital, London, UK

## Abstract

**Introduction:** Aquablation for surgical treatment of benign prostatic enlargement (BPE) causing bladder outflow obstruction (BOO) has demonstrated good functional outcomes, even for large glands, with high rates of ejaculatory preservation reported. This is a protocol for a study that aims to review real-world outcomes of ejaculatory preservation or restoration post-Aquablation in an unselected cohort and compare to published clinical trial outcomes.

**Methods:** Retrospective data will be collected from a prospectively maintained consecutive case series of patients who underwent Aquablation, in a single UK centre. The primary outcome is ejaculatory function subjectively reported by patients post-operatively, and classified as: antegrade ejaculation, retrograde/low volume ejaculation, anejaculation or not sexually active. Secondary outcomes are International Prostate Symptom Severity (IPSS), Quality of Life (QoL) Score, post-void residual (PVR), and incontinence. Descriptive and comparative statistical tests will be performed.

**Conclusions:** This is a protocol for a study to review real-world ejaculatory function and clinical outcomes following robotic Aquablation for prostatic bladder outflow obstruction and compare to published clinical trial outcomes.

## Introduction

Benign prostatic enlargement (BPE) is a major cause of bladder outflow obstruction (BOO) and has a significant impact on quality of life.^(1)^ Traditional surgical interventions include transurethral resection of the prostate (TURP) and laser enucleation. These often result in high rates of post-operative ejaculatory dysfunction, including retrograde (up to 75% in TURP, 90% in laser enucleation) and anejaculation, which is increasingly recognised as a key patient‒centred outcome.^(2)^

Robotic Aquablation is a waterjet‒based resection system integrating real‒time transrectal ultrasound guidance to achieve tissue removal.^(3)^ It is a relatively novel technique and is only starting to become used more widely in the UK. Clinical trials have demonstrated improvements in lower urinary tract symptoms (LUTS) and report high rates of ejaculatory function preservation, ranging from 72% to 99.6%.^(4,5,6,7,8)^

However, existing literature primarily reports outcomes from selected industry-sponsored trial cohorts. There is limited evidence from real-world unselected patient cohorts and few studies on post-operative ejaculatory function in men with pre-operative retrograde or low volume ejaculation.

## Study Aims and Objectives

The primary aim of this study is to describe ejaculatory function in men post-Aquablation. The secondary aims are to evaluate pre- and post-operative functional urinary outcomes, including: International Prostate Symptom Score (IPSS) and Quality of Life (QoL) score^(9)^, urinary flow rate (QMax) and post-void residual volume (PVR).

## Methods

### Study Design

This will be a retrospective observational cohort study using a prospectively maintained consecutive case series of patients who underwent Aquablation in a single UK tertiary Urology centre.

### Study Population

Inclusion criteria:

All male patients undergoing Aquablation for BPE-related BOO in the study period, with data available on or able to provide subjective assessment of sexual function and ejaculatory function at follow-up.

#### Exclusion criteria

Patients aged under 18; patients not sexually active and/or unable to comment on ejaculatory function.

### Study Outcomes

Primary outcome is rate of post-operative ejaculation preservation or restoration. Secondary outcomes are post-operative changes in functional urinary outcomes (IPSS and QoL score, urinary flow rate (QMax) and PVR) and subgroup comparisons (age, prostate volume, baseline characteristics).

### Data Collection and Analysis

Data will be extracted from electronic clinical records, including clinic letters, flow rate tests and patient-reported outcomes, from a prospectively maintained database. Where necessary, patients will be contacted via telephone to complete missing data not documented in electronic clinical records.

Data will be collected on the following variables: demographics (age), pre-operative data (prostate volume, IPSS, QoL, QMax, PVR, baseline ejaculatory function, urinary incontinence), operative details (resected weight, day case procedures, complications), post-operative data (IPSS, QoL, QMax, PVR, ejaculatory function, urinary incontinence, Patient Global Impression of Improvement (PGI-I)^(10)^) and time to follow-up.

Ejaculatory function will be subjectively assessed pre- and post-operatively and classified as: antegrade ejaculation, retrograde/low volume ejaculation, anejaculation or not sexually active.

Descriptive statistics will include mean, range and proportions. Comparative analysis will include Paired t‒tests or Wilcoxon signed-rank tests for pre‒ vs post‒operative outcomes, as appropriate. Subgroup analysis will be carried out based on baseline ejaculatory function, prostate size and age. A p-value of <0.05 will be considered statistically significant.

### Ethics and Dissemination

This study involves analysis of de-identified, routinely collected clinical data. It has been locally registered as a clinical audit. All data will be secured according to local governance procedures. This study will adhere to the STROBE (Strengthening the Reporting of Observational Studies in Epidemiology) guidelines and the STROBE Checklist will be made available in supplemental material of the final published study.

Results will be submitted for presentation at regional and national urological conferences and for publication in a peer-reviewed journal. The protocol will be available via MedRxiv prior to full manuscript submission.

All relevant data from this study will be made available by request upon study completion.

## Discussion

Aquablation is a relatively novel surgical technique in the management of BPE-related BOO. It is a particularly attractive option for patients focused on preserving ejaculatory function, however current literature is limited to selected trial cohorts.

This study will be useful to review a large real-world cohort and compare post-operative ejaculatory function and clinical outcomes following robotic Aquablation to published clinical trial outcomes. It will also include data on ejaculatory restoration outcomes, which remains an under-reported subset of patients.

This study will use subjective assessment of ejaculatory function to represent usual clinical care and will therefore represent real-world patient outcomes.

## Data Availability

All data produced in the present study are available upon reasonable request to the authors.

